# Predicting Early MASLD-HCC from Serum N-Glycomics: A SHAP-Interpreted Gaussian Naive Bayes Model Built on nLC-HCD-PRM-MS/MS Profiling

**DOI:** 10.64898/2026.08.04.26359485

**Authors:** Yu Lin, Chithravel Vadivalagan, Jianliang Dai, Suyu Liu, Natan Y. Lubman, David M. Lubman

## Abstract

Hepatocellular Carcinoma (HCC) arising from Metabolic Dysfunction-Associated Steatotic Liver Disease (MASLD) is an increasing public health burden with high mortality, highlighting the need for improved early detection strategies. Current surveillance tools, including Alpha-fetoprotein (AFP) and ultrasound, lack sufficient sensitivity for early-stage HCC detection. We analyzed serum samples from 131 patients, including 58 with cirrhosis and 73 with MASLD-related HCC (42 early-stage, 31 late-stage), using an nLC-stepped HCD-PRM-MS/MS workflow for targeted N-glycome profiling of glycopeptides derived from haptoglobin and vitronectin. Combining targeted glycopeptides with AFP significantly improved HCC detection compared with AFP alone. The optimal panel for all HCC versus cirrhosis (AFP + VTNC_169_A2G2F0S1 + VTNC_242_A3G3F2S2) achieved an AUC of 0.859 and 76.7% sensitivity at 90% specificity. For early-stage HCC, AFP + HP_184_A3G3F1S3 + VTNC_169_A2G2F0S1 yielded an AUC of 0.890 with 66.7% sensitivity at 1% specificity. A SHAP-selected Gaussian Naive Bayes model based on seven molecular/glycopeptide features, without demographic variables, further improved performance, achieving ROC-AUC values of 0.9985 in training and 1.0000 in independent testing cohorts, with accuracies of 98.1% and 100.0%, respectively.

**GRAPHICAL ABSTRACT:** 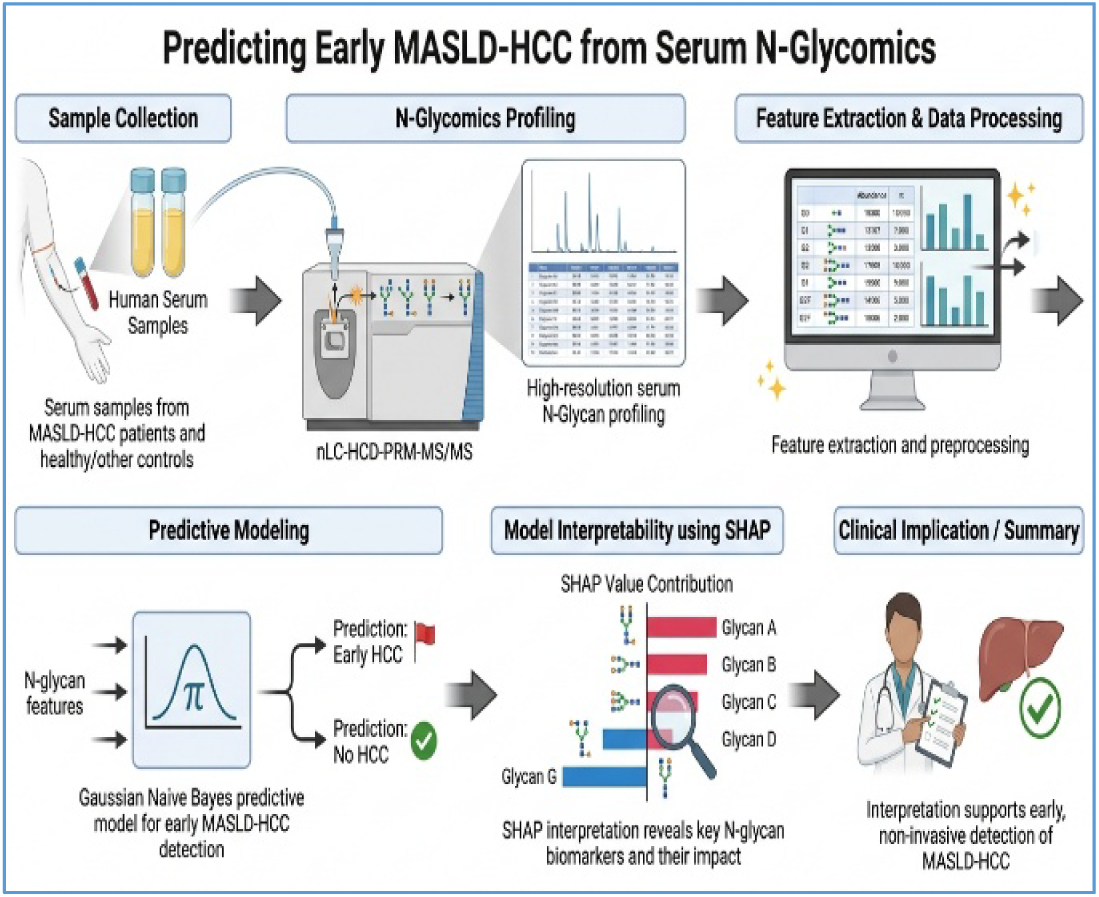

## 1. INTRODUCTION

Metabolic dysfunction-associated steatotic liver disease (MASLD), formerly termed nonalcoholic fatty liver disease (NAFLD), has emerged as a major etiological driver of hepatocellular carcinoma (HCC) and represents the fastest-growing contributor to liver cancer worldwide. Liver cancer accounts for more than 800,000 deaths annually, and the burden of MASLD-related HCC is increasing rapidly in parallel with the global rise in obesity and type 2 diabetes^1, 2^. Epidemiological projections suggest that MASLD may become the leading cause of liver cancer in several countries, underscoring its growing public health significance^3^. Early diagnosis of MASLD-HCC improves survival and clinical outcomes, highlighting the importance of routine HCC surveillance^4^.

Several diagnostic biomarkers currently used for HCC, including alpha-fetoprotein (AFP)^5^, glypican-3^6^, alpha-1-fucosidase^7^, and Golgi protein-73^8^, exhibit low sensitivity (∼60%), limiting their utility for early detection^9^. Therefore, improved and rigorously validated diagnostic alternatives are urgently needed. Post-translational modifications (PTMs) are essential regulatory mechanisms governing gene expression and protein function. Among these, glycosylation is a prominent PTM involving the covalent attachment of carbohydrate moieties (glycans) to proteins^10^. Glycosylation plays a critical role in protein folding, stability, and biological activity, and is fundamental to cellular communication processes such as cell adhesion and immune regulation^11^. This modification occurs on specific amino acid residues, predominantly asparagine in N-linked glycosylation and serine or threonine in O-linked glycosylation^12^.

Distinct glycosylation patterns contribute substantially to proteomic diversity, and their dysregulation has been strongly implicated in various pathological conditions, including tumor malignancy^11^. Sialyl Lewis (SLe) antigens are tumor-associated carbohydrate antigens that are frequently upregulated in several cancers, particularly pancreatic^13^, gastric^14^, and colorectal^15^. These antigens promote metastatic progression by meditating cancer cell adhesion to the vascular endothelium through binding to E-selectin^14^. Owing to their elevated expression in malignancies, SLe antigens serve as valuable diagnostic biomarkers for monitoring tumor progression^16^. Furthermore, their functional role in tumor cell adhesion and metastasis highlights their potential as promising therapeutic targets^17^.

Previous studies have shown that serum glycan structures associated with fucosylation and SLe antigens represent some of the most promising biomarkers for the early detection of HCC^18–21^. Targeted mass spectrometry-based approaches are particularly effective for the detection and quantification of well-established glycosylation and fucosylation sites in target proteins^22, 23^. Our recent research demonstrated that HCC associated with MASLD exhibits notable differences in fucosylated glycan structures^10^. Parallel Reaction Monitoring (PRM)-based target acquisition has demonstrated strong potential for glycan detection, providing deeper insights into biomarker discovery and enabling robust validation in larger cohorts, particularly through targeted capture and site-specific identification^10, 16, 24–27^.

Protein digestion is a critical step in glycoproteomic analysis, particularly for target-based acquisition, which significantly influences glycan structure identification and quantification^28^. In most of our previous studies, analyses were performed on a single target protein using a double-digestion strategy (Trypsin/Glu-C), which substantially enhanced the detection and characterization of glycan structures^10, 29^. However, this approach may incur significant costs and require substantial time. To address these issues, we need a low-cost, time-efficient glycoproteomic approach that can reliably identify early biomarkers for HCC.

The present study addresses the limitations of AFP by exploring effective and low-cost diagnostic approaches for MASLD-associated HCC. We analyzed whole serum samples from a large cohort of MASLD-associated HCC patients. A single-digestion workflow was employed, followed by targeted PRM analysis focusing on two key proteins, haptoglobin (Hp) and vitronectin (VTNC). Previously, we applied data-dependent acquisition (DDA), which involves random precursor (m/z) selection from pooled serum samples, and subsequently used PRM for the quantification of selected target proteins. The primary objective of this study was to thoroughly evaluate the diagnostic performance of a glycopeptide panel, both individually and in combination with AFP and other established biomarkers. In addition, selected glycan features and complementary markers were integrated into machine-learning models to develop robust biomarkers for the early detection of MASLD-associated HCC. To complement the conventional ROC and logistic-regression analyses, we further evaluated a compact machine-learning classifier based on seven SHAP-selected molecular features. Importantly, the model deliberately excluded demographic variables, reducing the risk that classification performance would depend on cohort-specific age or sex distributions and simplifying potential prospective clinical-trial implementation.

## 2. MATERIALS AND METHODS

### 2.1. Materials

Reagents were purchased from Sigma unless otherwise specified. Sequencing-grade trypsin was obtained from Promega (Madison, WI). Zeba Spin Desalting columns (7 kDa MWCO) were purchased from Thermo Scientific (Rockford, IL). The HILIC-packed TopTips were obtained from Glygen (Columbia, MD).

### 2.2. Serum samples

Serum samples were obtained from patients with MASLD cirrhosis (n = 58), early-stage MASLD associated HCC (n = 42), and late-stage MASLD associated HCC (n = 31) were treated at Shenzhen Hospital. MASLD associated cirrhosis and HCC were clinically defined by the presence of the metabolic syndrome components (including obesity, diabetes mellitus, and dyslipidemia) and the absence of other etiologies of chronic liver disease such as viral hepatitis or alcohol abuse. The clinical features of patients are summarized in **Table 1**. Samples were aliquoted and stored at -80 °C without undergoing any prior thaw cycles. The 131 patient serum samples were categorized as two groups: cirrhosis (n = 58) and MASLD-associated HCC (n = 72), including early-stage (n = 42) and late-stage (n = 31) disease. Early-stage HCC was defined according to the Milan criteria, the most widely used criteria for liver transplantation in People’s Republic of China (PRC). Institutional Review Board (IRB) approval for the study was obtained from Shenzhen Hospital.

**Table 1.**
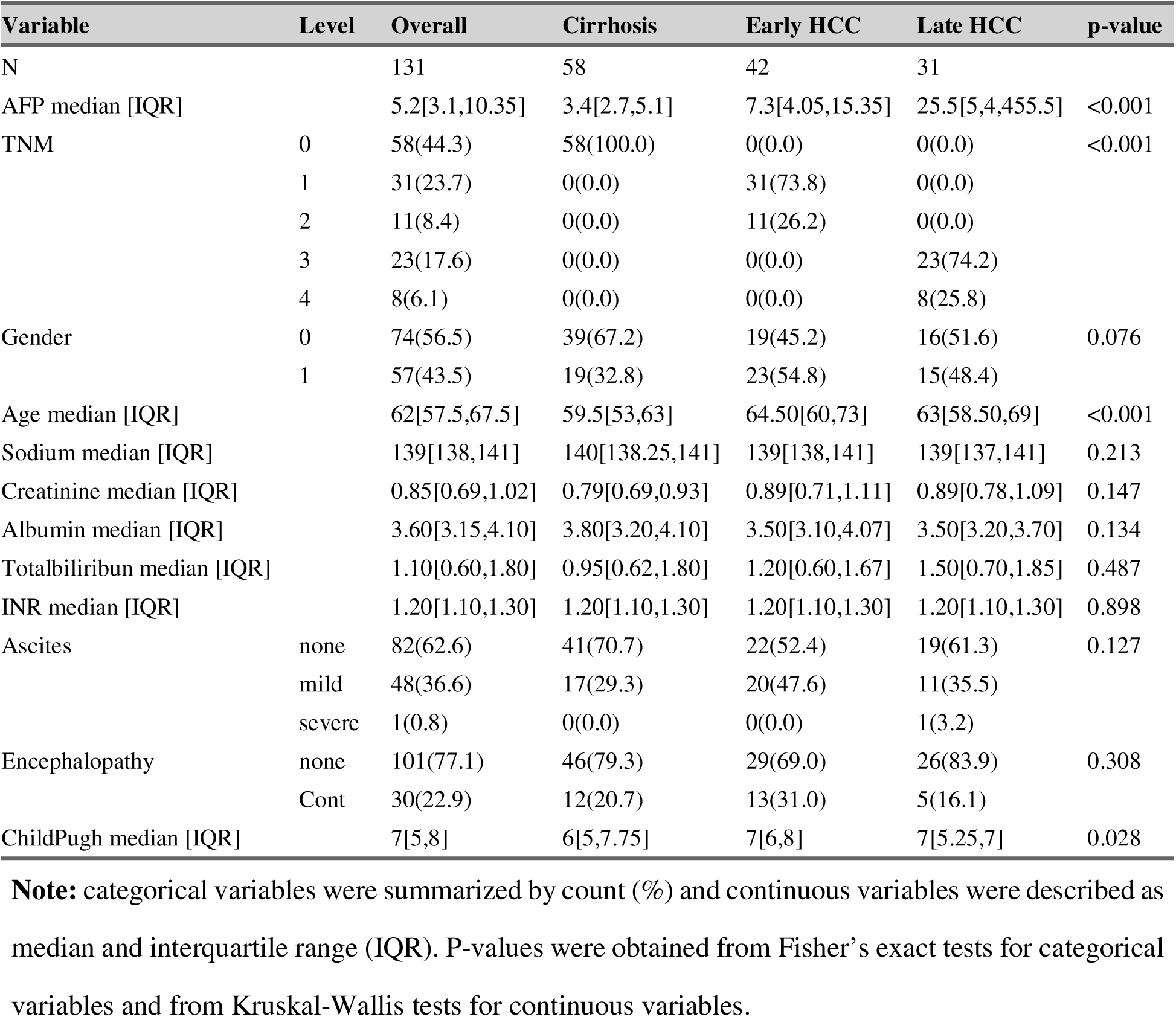
Summary of Clinical Characteristics of Patients.

**Table 2:** Performance of the SHAP-selected Gaussian Naive Bayes model derived from the ROC curves and confusion matrices.

| *Cohort* | *n* | *ROC/AUC* | *Accuracy* | *Sensitivity* | *Specificity* | *Accuracy* | *F1-score* | *Confusion matrix* |
| --- | --- | --- | --- | --- | --- | --- | --- | --- |
| *Training* | 105 | 0.9985 | 98.1% | 96.6% | 100.0% | 98.3% | 98.2% | TN=47, FP=0, FN=2, TP=56 |
| *Testing* | 26 | 1.0000 | 100.0% | 100.0% | 100.0% | 100.0% | 100.0% | TN=11, FP=0, FN=0, TP=15 |

**Table 3:** Exploratory comparison of the SHAP-selected Gaussian Naive Bayes classifier with AFP alone, logistic-regression biomarker panels from the present manuscript, and related glycopeptide-based HCC biomarker approaches.

| *Method / model* | *Comparison* | *AUC* | *Accuracy* | *Sensitivity / specificity* | *Interpretation* |
| --- | --- | --- | --- | --- | --- |
| *AFP alone* | All HCC vs cirrhosis | 0.788 | Not reported | 67.9% / 80% | Baseline clinical biomarker |
| *AFP alone* | Early HCC vs cirrhosis | 0.768 | Not reported | 66.7% / 80% | Limited early-stage sensitivity |
| *AFP + N241_A4G4F1S3* | All HCC vs cirrhosis | 0.834 | Not reported | 65.8% / 85% s | Best reported two-marker Hp panel for all-stage HCC |
| *AFP + HP_184_A3G3F1S3* | Early HCC vs cirrhosis | 0.872 | Not reported | Not reported in text | Strong two-marker early-stage panel |
| *AFP + VTNC_169_A2G1F0S1 + VTNC_242_A3G3F2S2* | All HCC vs cirrhosis | 0.859 | Not reported | 78.1% / 85% | Best three-marker all-stage panel in this manuscript |
| *AFP + Age + HP_184_A3G3F1S3* | Early HCC vs cirrhosis | 0.904 | Not reported | 81.5% / 85% | Best early-stage panel in this manuscript; includes age |
| *Prior Hp Sialyl Lewis glycopeptide + AFP panel* | NASH/MASLD-HCC vs cirrhosis | 0.898 | Not reported | Not reported in this manuscript section | Previously reported glycopeptide-based panel |
| *SHAP-selected Gaussian Naive Bayes* | Training cohort | 0.9985 | 98.1% | 96.6% / 100.0% | Near-perfect internal training performance; no demographic variables used |
| *SHAP-selected Gaussian Naive Bayes* | Independent testing cohort | 1.0000 | 100.0% | 100.0% / 100.0% | Near-perfect held-out testing performance; no demographic variables used |

**Table 4:**
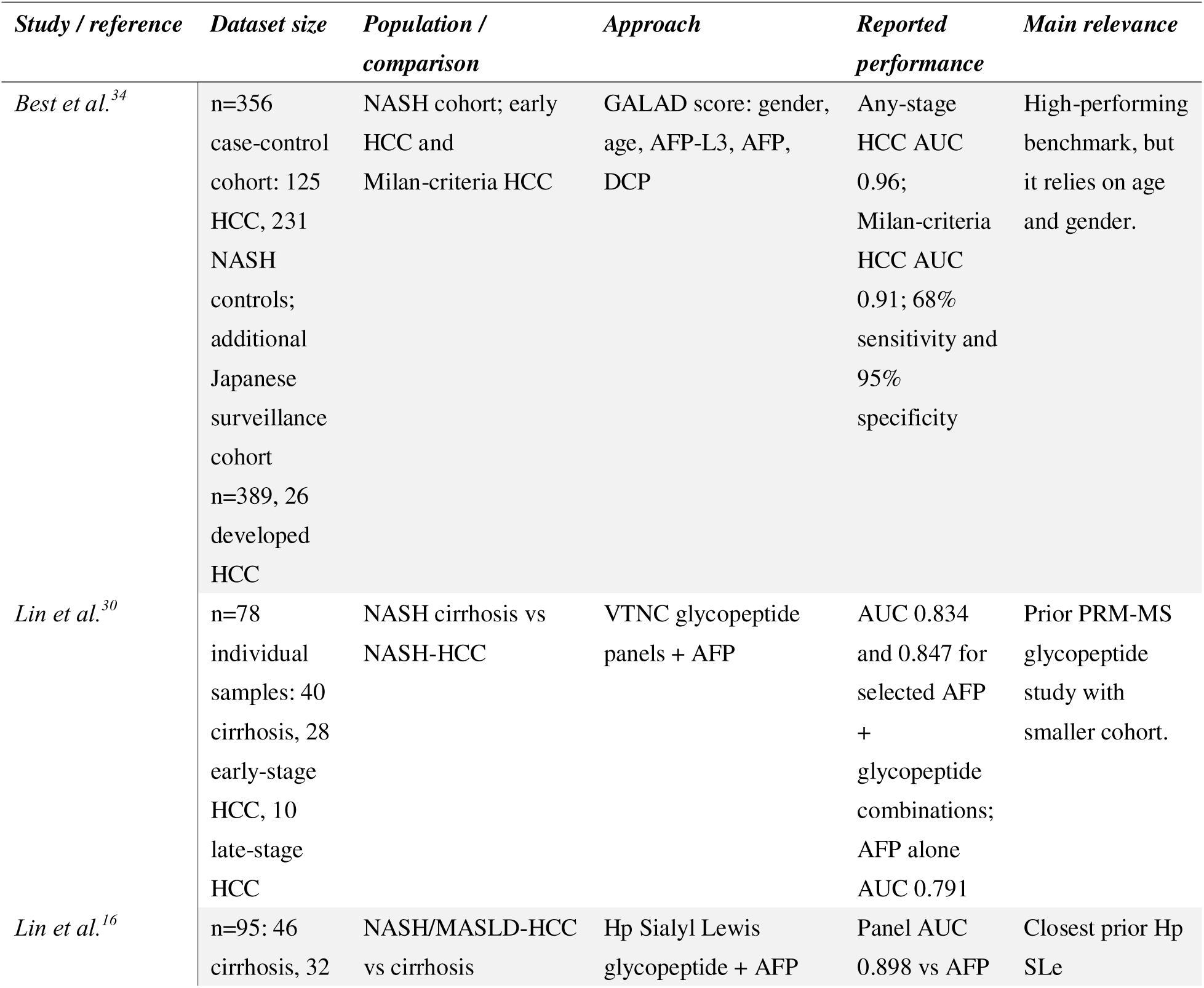
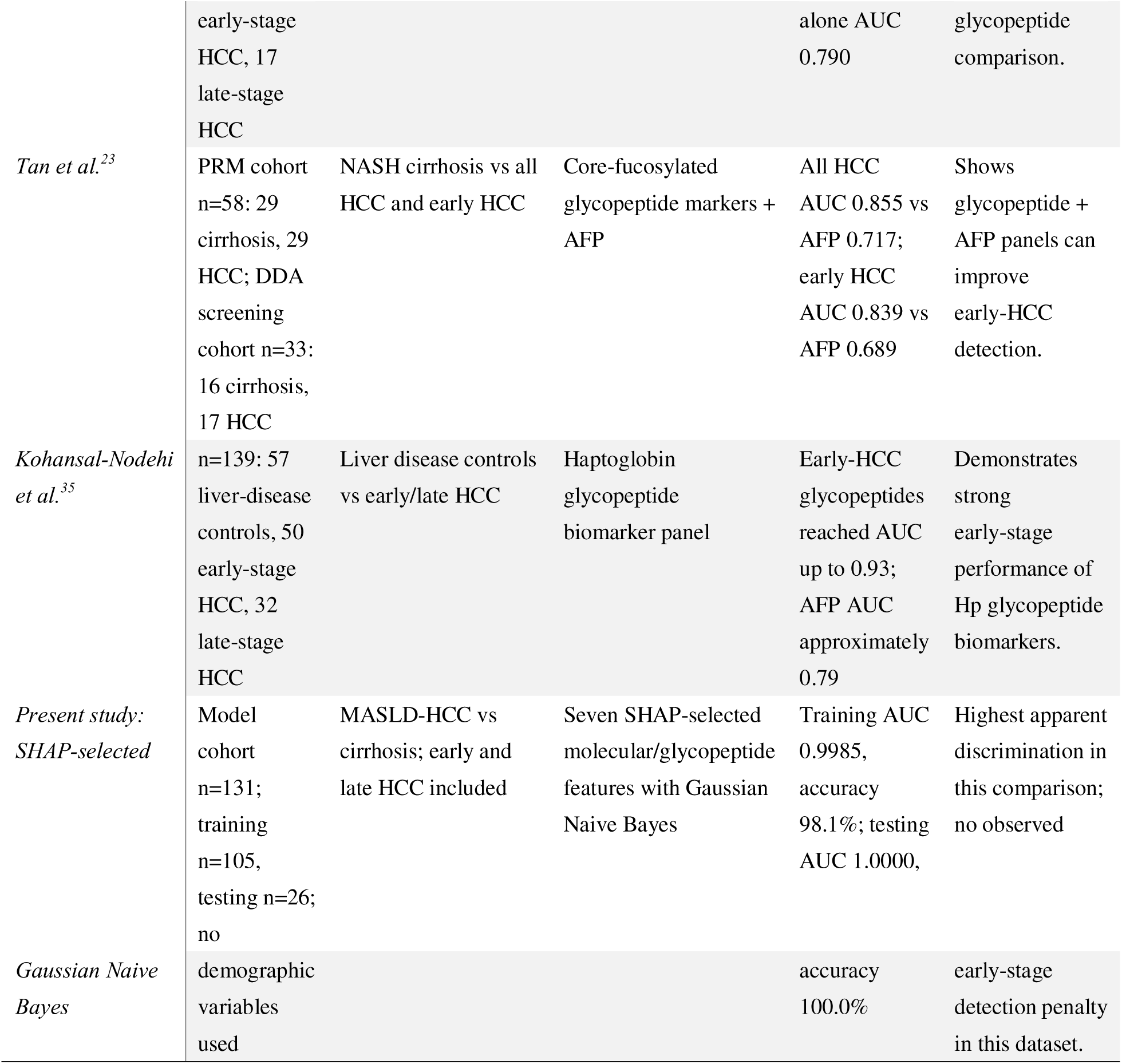
Comparison with selected published HCC detection studies and biomarker approaches.

### 2.3. Serum Glycopeptide Enrichment for LC-Stepped HCD-DDA-MS/MS

The workflow for the initial screening of glycopeptides is illustrated in Figure 1. Pooled serum samples were prepared by combining 10 µL of serum from each of 10 patients per group, including cirrhosis, early-stage MASLD HCC, and late-stage MASLD HCC. For each pooled sample, 10 μL of serum was diluted with 200 μL PBS and filtered through a 0.22 μm membrane to remove the debris. The filtered serum samples were subsequently desalted using an Ultra-4 centrifugal filter (10 kDa MWCO, Millipore) and dried using a SpeedVac concentrator (Thermo).

**Figure 1.**
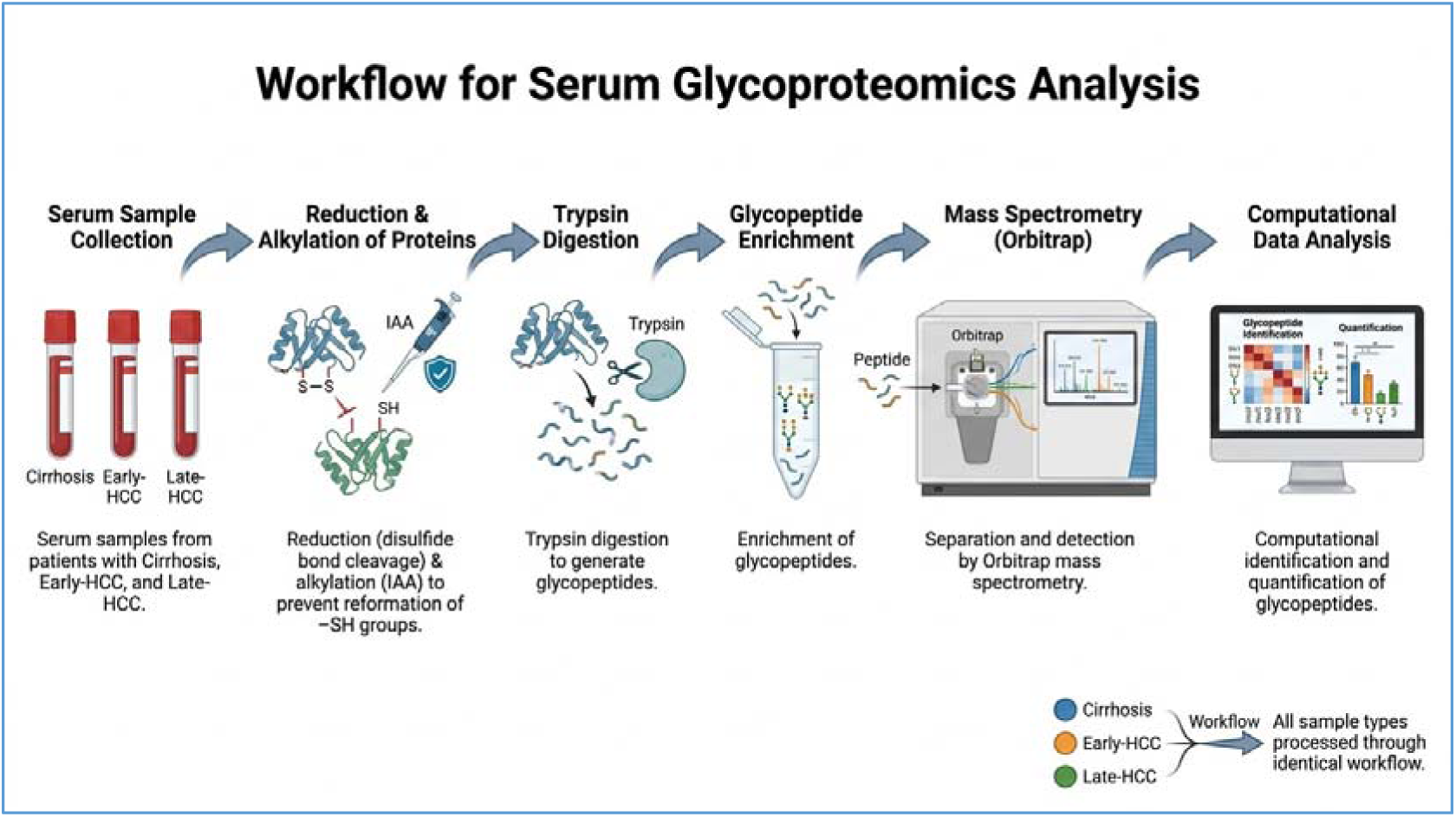
Workflow for serum glycoproteomics analysis. Individual serum samples from patients with cirrhosis, Early-HCC, and Late-HCC were collected. Proteins in the serum were first reduced by breaking disulfide bonds and IAA to prevent reformation of -SH groups. The reduced proteins were then digested with trypsin to generate glycopeptides. Glycopeptides were subsequently enriched and analyzed using mass spectrometry on an Orbitrap instrument, where they were separated and detected. Finally, the resulting mass spectrometry data were subjected to computational analysis to identify and quantify glycopeptides.

The dried protein sample was reconstituted in 50 mM ammonium bicarbonate (pH 8.2). Disulfide bonds were reduced by adding dithiothreitol (DTT) to a final concentration of 20 mM and incubating at 60 °C for 30 min. Alkylation of free thiol groups was performed by adding iodoacetamide (IAA) to a final concentration of 50 mM and incubating for 30 min at room temperature in the dark. Excess IAA was quenched by the addition of DTT to a final concentration of 5 mM. The samples were desalted using a Zeba Spin desalting column (Thermo Scientific) and dried. The desalted sample was then redissolved in 50 mM ammonium bicarbonate, followed by the addition of 3μL of trypsin (400 ng/μL, Promega, Madison, WI) and incubated at 37 °C for 14 h. After digestion, the sample was dried in a SpeedVac concentrator (Thermo) prior to glycopeptide enrichment.

Glycopeptides were enriched using a HILIC TopTip (Glygen, Columbia, MD) as previously described^23^. Briefly, the tryptic digest was reconstituted in binding buffer (15 mM ammonium acetate with 85% acetonitrile, pH 3.5) and loaded onto a pre-equilibrated HILIC tip to allow glycopeptides binding. The tip was washed three times with binding buffer to remove non-bound peptides. Glycopeptides were then eluted with water and dried using a SpeedVac concentrator (Thermo).

### 2.4. Glycopeptides Enrichment by Filter for LC-Stepped HCD-PRM-MS/MS

The digested glycopeptides used for LC-Stepped HCD-PRM-MS/MS analysis were prepared from a total of 131 individual serum samples as described above. Briefly, each serum sample was reduced with DTT, alkylated with IAA, and subjected to tryptic digestion. Following digestion, glycopeptides were enriched by a 3 kDa filter and further purified by buffer exchange with distilled water performed five times via centrifugation. The resulting glycopeptide solutions were subsequently dried using a SpeedVac concentrator. The enriched glycopeptides were further analyzed by mass spectrometry to quantify glycoprotein expression and glycosylation patterns.

### 2.5. LC-Stepped HCD-DDA-MS/MS

The dried glycopeptide fractions were reconstituted in 0.1% formic acid (FA) and analyzed in duplicate on an Orbitrap Fusion Lumos Tribrid Mass Spectrometer (Thermo Fisher Scientific) coupled to a Dionex UPLC system. Peptides were separated using a binary solvent system consisting of 0.1% FA in H_2_O (solvent A) and 80% acetonitrile containing 0.1% FA (solvent B). Separation was performed on a 75 μm × 50 cm C18 column (2 μm, 100 Å; Acclaim PepMap RSLC, Thermo Fisher Scientific) using a 90 min linear gradient from 2 to 40% solvent B at a flow rate of 300 nL/min. The MS instrument was operated in data dependent mode. Full MS1 scans (m/z 400–1800) were acquired in the Orbitrap at a resolution of 120,000 with an AGC target of 4 × 10 and a maximum injection time of 100 ms. The most highly charged precursor ions were selected in order of intensity for stepped higher-energy collisional dissociation (HCD) MS/MS, with fragment ions detected in the Orbitrap at a resolution of 60,000 using an AGC target of 2 × 10 and a maximum injection time of 250 ms. Stepped HCD collision energies of 31.5%, 35%, and 38.5% were applied, as optimized for large-scale glycopeptide characterization in this study.

### 2.6. LC-Stepped-HCD-PRM-MS/MS

The LC-MS system was identical to that used for the DDA detection mode described above. Peptides were separated using a 65-min linear gradient of 2-40% solvent B at a flow rate of 300 nL/min. For PRM analysis, stepped collision energies (CEs) of 19%, 26%, and 33% were applied to fragment glycopeptides, differing from the settings used in DDA mode. Unlike DDA, which scans all precursor ions, PRM analysis relies on predefined precursor ions, resulting in improved detection sensitivity. In the present study, glycopeptides were selected as targeted precursor ions, and the peptide backbone containing a single HexNAc moiety (pep+HexNAc) was defined as Y1 ion for quantitative analysis. The targeted precursor ions originated from two proteins, including Hp and VTNC. To determine the retention time of the targeted precursors, an initial survey scan was performed in DDA mode prior to PRM analysis.

### 2.7. Data Interpretation and Relative Quantitation

For the DDA results, all spectra were searched with pGlyco 3.1, a software for peptide, glycopeptide and protein identification based on tandem MS spectra, as described previously. A UniProt human protein database containing 20,359 proteins was used for data searching. The search was performed with the following parameters: (1) fixed modification, carbamidomethyl (C); (2) variable modifications, oxidation (M); deamidation (N, Q) and N-glycan modifications (N); (3) up to one missed cleavage; and (4) mass tolerance of 10 ppm for MS1 and 20 ppm for MS2. The theoretical m/z values of the oxonium ions derived from glycan fragments in HCD-MS were used for glycan identification, including GlcNAc (m/z 138.05, m/z 168.05, and m/z 204.09), NeuAc (m/z 274.09 and 292.10), GlcNAc-Hex (m/z 366.14), HexHexNAcFuc (m/z 512.20), and HexNAcHexNeuAc (m/ z 657.23).

Automatic quantitative analysis was performed using Skyline software as described previously. The peak area of the extracted ion chromatogram (XIC) for each glycopeptide was integrated and normalized to the sum of peak areas of all glycopeptides identified within the same MS run, yielding the relative abundance of each N-glycopeptide in the sample. The abundance of a site-specific glycoform was calculated as the sum of the glycopeptides carrying the same glycan at the corresponding glycosylation site.

For the PRM data analysis, Skyline software was used for the quantification of the selected glycopeptides. Glycopeptide identification was based on characteristic oxonium ions, including GlcNAc, NeuAc, GlcNAc-Hex, HexHexNAcFuc, and HexNAcHexNeuAc as well as other relevant b/y ions. Quantification was performed using the Y1 ion (peptide + HexNAc). A spectral library was generated using parameters obtained from a survey scan acquired prior to PRM analysis, including glycopeptide sequence, scan number, retention time, charges state, and a.ms2 file converted from the survey scan raw data. A FASTA file containing the targeted proteins was used as the background protein database. For transition settings, precursor charge states were set from +2 to +5, fragment ion charge states were set to +1 and +2, and ion types were limited to b and y ions. The ion match tolerance was set as 0.05 m/z. Y1 ions corresponding to the targeted glycopeptides were manually defined as (target peptide+HexNAc). The integral peak areas of the Y1 ions were exported from Skyline in .csv format and used for glycopeptide quantification and data normalization. Relative glycopeptide abundance was calculated by normalizing the peak area of each glycopeptide to the sum peak areas of all targeted glycopeptides within each sample. Relative abundances were then compared across different disease-state groups, and scatter plots were generated using GraphPad Prism software (version 9.0).

### 2.8. Statistical analysis

Descriptive statistics were used to summarize patient characteristics. Group differences were assessed using Fisher’s exact test for categorical variables (e.g., sex) and the Kruskal−Wallis test for continuous variables (e.g., age). Marker distributions were summarized using descriptive statistics, including the median and the range, and were visualized using histogram. The Wilcoxon rank-sum test was used to compare marker levels between HCC and cirrhosis samples. P-values were adjusted for multiple comparisons using the Bonferroni correction. Markers that were clinically relevant and showed statistically significant differences between HCC and cirrhosis were selected as the candidate markers for panel development. Their differentiation performance was evaluated using the area under the curve (AUC) from receiver operation characteristic (ROC) analysis. The logistic regression model was used to combine the site-specific glycopeptide biomarker candidates with AFP during panel development. The optimal two-marker and three-marker panels were selected based on their estimated AUC values. Bootstrapping was used to compare the AUC of the selected panel with that of AFP alone by testing the hypothesis H : AUC_panel = AUC_AFP versus H : AUC_panel ≠ AUC_AFP. A two-sided P value < 0.05 was considered statistically significant. Youden’s index was used to determine the optimal cutoff by maximizing the combined sensitivity and specificity of the continuous variable. All statistical analyses were performed using R Statistical Software (version 4.4.1; R Foundation for Statistical Computing, Vienna, Austria).

### 2.9. Machine Learning Analysis

A machine learning framework was implemented to evaluate the diagnostic potential of selected glycopeptide biomarkers for distinguishing MASLD-associated HCC from cirrhosis controls. Seven features were selected using SHAP (SHapley Additive exPlanations)-based feature importance analysis and were used to train a Gaussian Naive Bayes classifier (HP_241_ A3G3F1S3; A4G4F1S3; A2G2F1S2; VTNC_242_ A3G3F2S2; A3G3F2S3; A2G2F0S1; A3G3F1S3.

No demographic variables were included in the Gaussian Naive Bayes model. Specifically, age, sex/gender, ethnicity, and other cohort-descriptive clinical variables were excluded from model input. This was done intentionally because demographic variables can inflate apparent diagnostic performance in small or imbalanced datasets and may complicate prospective trial design by requiring demographic matching, stratification, or adjustment. The resulting model therefore reflects molecular/glycopeptide information rather than demographic separation between cohorts.

The dataset was separated at the patient level into independent training and testing cohorts. Based on the confusion matrices, the training cohort included 105 patients (80.2% of the model cohort), including 47 cirrhosis controls and 58 HCC cases, whereas the testing cohort included 26 patients (19.8% of the model cohort), including 11 cirrhosis controls and 15 HCC cases. Thus, the effective split was approximately 80.2%/19.8% train/test.

In datasets of this size, k-fold cross-validation is commonly used because retaining an independent test set may reduce the number of samples available for model training. In the present analysis, however, the priori expectation for strong discrimination was high and the selected features showed very strong separation. This allowed a held-out patient-level testing cohort to be preserved without substantially compromising training stability. The held-out design provides a more clinically interpretable estimate of model generalization than reporting only cross-validated performance. The patient-level separation was performed before model assessment. SHAP-based feature selection should be reported as being performed exclusively within the training cohort to minimize information leakage.

Gaussian Naive Bayes was selected because it is appropriate for continuous biomarker abundance values, provides a parsimonious probabilistic model, and is less prone to overfitting than highly flexible machine-learning algorithms in modest-sized clinical cohorts. Model performance was evaluated using ROC-AUC, confusion matrices, accuracy, sensitivity, specificity, balanced accuracy, precision, and F1-score. Accuracy was calculated as (TP + TN)/(TP + TN + FP + FN), sensitivity as TP/(TP + FN), specificity as TN/(TN + FP), precision as TP/(TP + FP), balanced accuracy as the mean of sensitivity and specificity, and F1-score as 2 x precision x sensitivity/(precision + sensitivity).

## 3. RESULTS AND DISCUSSION

### 3.1. Classification of Serum Samples Based on Patient Characteristics

A total of 131 patients were included in the study, containing 58 with MASLD cirrhosis, 42 with early MASLD HCC, and 31 with late MASLD HCC. Significant differences were observed among the three groups in AFP levels, TNM stage, gender distribution, age, and Child-Pugh score. AFP levels progressively increased from the cirrhosis group to early and late MASLD HCC (p < 0.001). TNM stage distribution varied as expected, with cirrhosis patients classified as stage 0, early MASLD HCC as stages 1-2, and late MASLD HCC as stages 3-4 (p < 0.001). Age differed modestly across groups (p = 0.003), with patients in the early MASLD HCC group being slightly older. Gender distribution also showed a significant difference among the groups (p = 0.032). The Child-Pugh score was higher in the early MASLD HCC group compared with the cirrhosis and late MASLD HCC groups (p = 0.008). In contrast, no significant differences were observed in serum sodium, creatinine, albumin, total bilirubin, INR, ascites, or hepatic encephalopathy among the groups. Serum samples were categorized as MASLD cirrhosis or MASLD HCC based on the clinical characteristics summarized in Table 1.

### 3.2. Glycopeptide Biomarker Candidates Identified using LC-Stepped-HCD-PRM-MS/MS

To validate the differential expression of glycopeptides and assess their diagnostic potential for detection of MASLD-associated HCC, LC-stepped-HCD-PRM-MS/MS was performed for targeted quantitative analysis of the selected glycopeptide candidates across distinct liver disease states, including 49 cases of cirrhosis, 33 cases of early-stage MASLD HCC, and 25 cases of late-stage MASLD HCC. Targeted glycopeptides for PRM-MS analysis were based on our previous report identifying site-specific glycopeptides of Hp and VTNC that exhibited significant alterations during the progression from MASLD cirrhosis to MASLD HCC as determined by differential LC-DDA-MS/MS analysis^30^. The candidate glycopeptide markers were predominantly characterized by fucosylated and/or highly sialylated glycan motifs. Although the overall level of serum Hp fucosylation can be measured using a lectin antibody enzyme-linked immunosorbent assay (ELISA), this approach does not capture the subtle yet biologically significant changes in specific Hp glycoforms.

PRM-MS offers a significant advantage over DDA-MS in that it enables targeted quantitative analysis, whereas DDA-MS scans across the entire mass range^10^. The enhanced sensitivity of PRM arises from its targeted acquisition strategy, which allows the mass spectrometer to allocate longer ion accumulation times to selected precursor ions (i.e., increased dwell time), compared with DDA-MS^31^. Moreover, PRM enables the parallel measurement of all fragment ions derived from a predefined precursor allowing acquisition of a full MS2 product ion spectrum for a specified precursor ion. Consequently, all detectable product ions can be monitored simultaneously with high mass accuracy and resolving power. In addition, the PRM-MS provides a broad dynamic range for quantitative analysis of target precursors^32^. Collectively, these features facilitate accurate quantification of a larger number of predefined precursors within an expected chromatographic elution window.

In a recent study, four glycopeptides site N207 from Hp were reported to exhibit significantly altered abundance between HCC and cirrhosis in serum samples from a cohort of 30 patients. Furthermore, the combination of AFP with glycopeptides N207_A3G3F1S2 and N207_A3G3F1S3 as a three-marker panel, demonstrated improved diagnostic performance compared with AFP alone, yielding a higher AUC^33^. In another study^16^, glycopeptides bearing SLe antigens in Serum Hp were identified as candidate biomarkers for nonalcoholic steatohepatitis (NASH), especially the tetra-antennary glycoform of glycopeptides, when combined with AFP, significantly improve diagnostic accuracy, achieving an AUC of 0.898 (95% CI: 0.835, 0.951), compared with an AUC of 0.790 (95% CI, 0.697 0.872) for AFP alone (P = 0.048).

In the present study, the sample cohort comprised 131 patient serum samples, representing an expanded dataset. The precursor ions selected for the current experiment were based on previous work, in which both +3 and +4 charge states were included^10^. These methodological differences enabled improved accuracy and precision, revealing significant changes in glycopeptides abundance at sites N184 and N241 from Hp, as well as N169 and N242 from VTNC. Furthermore, in contrast to our prior study, the target proteins were not physically isolated prior to analysis, rather PRM was performed directly on the total protein mixture obtained from whole serum.

In the present study, 49 precursor ions derived from site-specific glycopeptides were selected for PRM-MS analysis across different liver disease states. These glycopeptides included mono- and bi-fucosylated glycoforms at sites N184, and N241 of Hp, as well as N169 and N242 of VTNC. For selected glycopeptides, both +3 and +4 charge states of the precursor ion were included in the PRM-MS analysis based on MS profiles obtained in DDA mode. Quantitative analysis was performed using the Skyline platform to determine the relative abundance of each glycopeptide. Five Hp-derived glycopeptides and Five VTNC derived glycopeptides exhibited statistically significant differences between patients with HCC and those with cirrhosis (Figure 2 & 3). The relative abundance of each glycopeptide was normalized to its corresponding peak intensity, and the normalized value was used for the ROC analysis.

**Figure 2.1:**
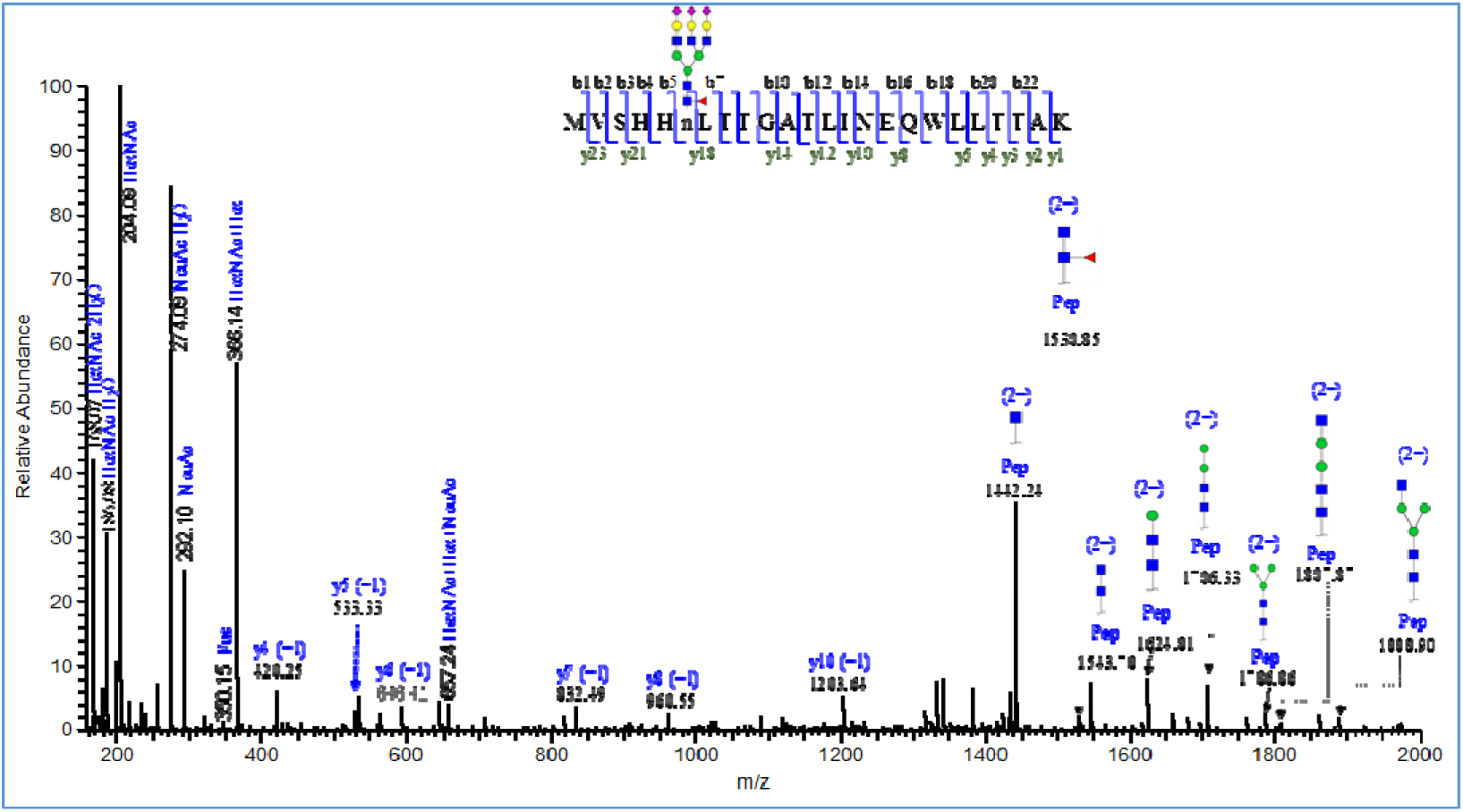
A representative tandem mass spectrum of glycopeptide of HP_N184_MVSHHnLTTGATLINEQWLLTTAK HexNAc(5)Hex(6)Fuc(1)NeuAc(1)_A3G3F1S3

**Figure 2.1:**
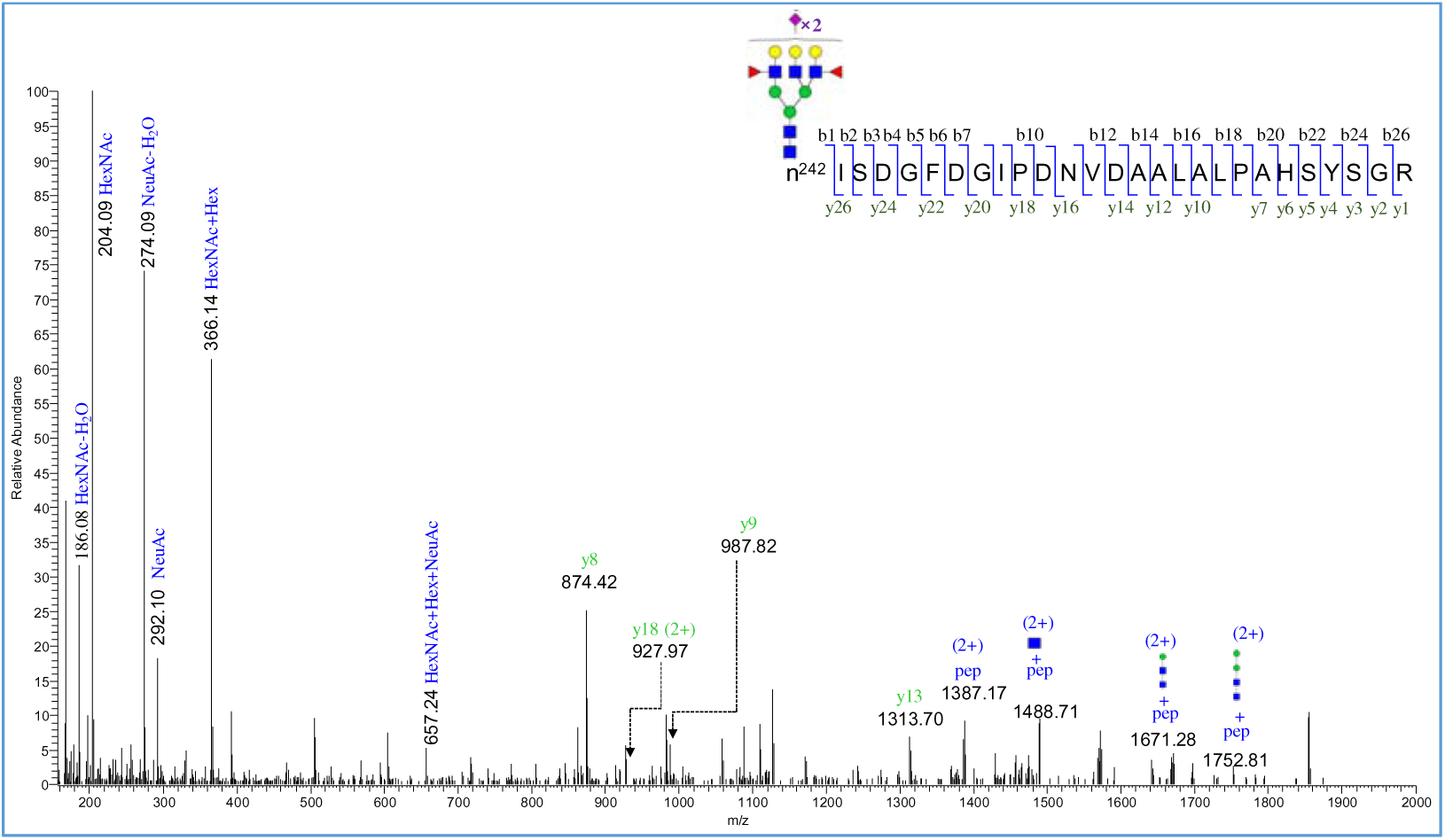
A representative tandem mass spectrum of glycopeptide from VTNC of N242 n^242^ISDGFDGIPDNVDAALALPAHSYSGR _A3G3F1S2

**Figure 2.2:**
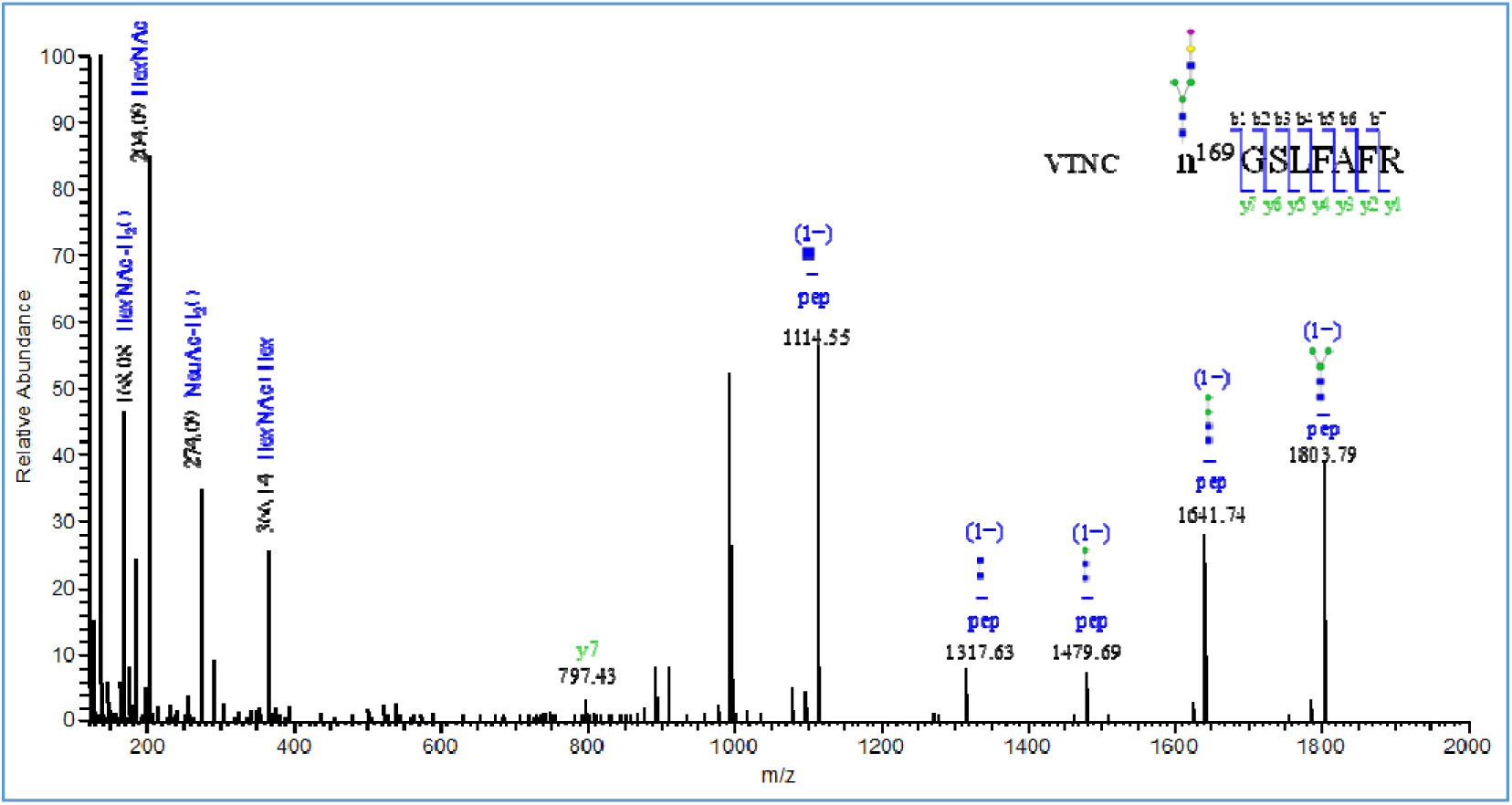
A representative tandem mass spectrum of glycopeptide from VTNC_N169_ n169 GSLFAFR_ A2G2F0S1.

**Figure 3.**
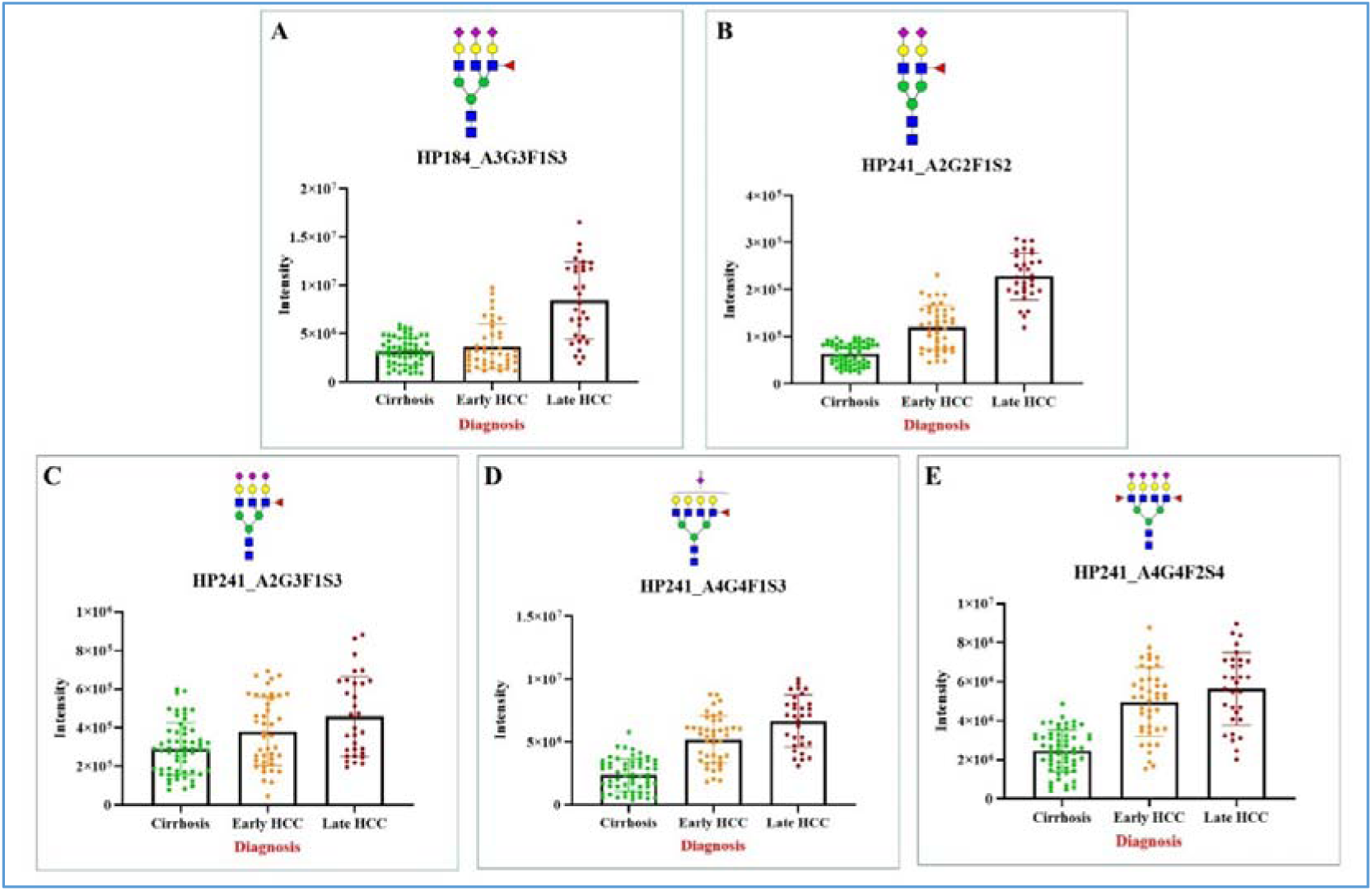
Relative abundance of site-specific Hp glycopeptides across disease stages. Scatter plots with overlaid bar graphs depict the intensity levels of individual glycopeptides in patients with cirrhosis, early HCC, and late HCC. Each dot represents an individual patient sample, while bars indicate the mean ± SD. Representative glycan structures are shown above each corresponding panel. (A) HP184_A3G3F1S3; (B) HP241_A2G2F1S2; (C) HP241_A2G3F1S3; (D) HP241_A4G4F1S3; (E) HP241_A4G4F2S4.

Among the five significant Hp glycopeptides, two glycosylation sites were involved including N184 and N241. At site N184, the glycopeptide N184_ A2G2F1S3 was identified. Four glycopeptides derived from the same peptide backbone containing N241 were detected including the bi-antennary glycoform N241_A2G2F1S2, tri-antennary glycoform N241_A3G3F1S3, tetra-antennary glycoform N241_A4G4F1S3, and N241_A4G4F2S4. Five significant VTNC glycopeptides were identified across two glycosylation sites, N169 and N242. Among these, the bi-antennary glycoforms N169_A2G2F0S1, N242_A2G2F0S2, and N242_A2G2F0S1 were detected. In addition, the tri-antennary glycoforms N242_A3G3F2S2, N242_A3G3F2S3, and N242_A3G3F1S3 were also detected.

All detected fucosylated and Sialylated glycoforms from Hp and VTNC were elevated in HCC compared with cirrhosis, highlighting the increased complexity of fucosylation and sialylation in HCC, consistent with previous reports^30^. Notably, these five Hp glycopeptides also demonstrated statistically significant differences between patients with early-stage HCC and those with cirrhosis. Importantly, oxonium ions at m/z 657.24 (HexNAc-Hex-NeuAc) were clearly observed in the MS/MS spectra of these five glycopeptides, indicating that these peptides are sialylated glycopeptides. Among the Hp-derived glycopeptides, the mass spectrum of glycopeptide N184_A3G3F1S3 (m/z 1530.85; peptide + HexNAc + HexNAc + Fuc) revealed a core-fucosylated glycoform (Figure-2.1&2.2). For VTNC, five glycopeptides were identified. Among these, the sialylated bi-antennary glycopeptide at glycosylation site N169_A2G2F0S1 exhibited significant differences among the study groups (Figure 2.3).

### 3.3. Diagnostic Performance of Site-Specific N-Glycopeptides in Overall and Early-Stage HCC

ROC analysis was performed to evaluate the performance of Hp site-specific N-glycopeptides in distinguishing all HCC cases and early HCC cases from cirrhosis controls. The estimated AUC values and corresponding 95% confidence interval (CI) for each individual marker are summarized in Table S4. When comparing all HCC cases with cirrhosis, AFP demonstrated an AUC of 0.788 (95% CI: 0.711, 0.861), with a sensitivity of 67.9 at 80% specificity. The five glycopeptides derived from two Hp glycosites (N184 and N241) which were significantly differentially expressed between cirrhosis and HCC as described above, were further evaluated (Figure-3). The glycopeptide involving site N184 yielded an AUC of 0.637 (95% CI 0.528,0.731). Four glycopeptides were derived from glycosite N241, including one bi-antennary glycopeptide (N241_A2G2F1S2), one tri-antennary glycopeptide (N241_A3G3F1S3), and two tetra-antennary glycopeptides (N241_A4G4F1S3 and N241_A4G4F2S4). The corresponding AUCs were 0.650 (95% CI:0.551,0.746) for N241_A2G2F1S2, 0.672(0.582,0.756) for N241_A3G3F1S3, 0.662(0.565,0.754) for N241_A4G4F1S3 and 0.552 (0.444,0.656) for N241_A4G4F2S4.

For the comparison between early-stage HCCs to cirrhosis, AFP achieved an AUC of 0.768 (95% CI:0.672,0.86) with a sensitivity of 66.7 at 80% specificity. Consistent with the findings in the overall HCC cohort, glycopeptides involving the N184 site did not outperform AFP alone. Among the N241 derived glycopeptides, the AUCs were 0.562 (0.455,0.668) for N241_A2G2F1S2, 0.624 (0.514,0.725) for N241_ A3G3F1S3, 0.715 (0.609,0.814) for N241_ A4G4F1S3 and 0.620 (0.509,0.723) for N241_ A4G4F2S4. Overall, AFP demonstrated superior discriminatory performance compared with individual Hp site-specific glycopeptides, although certain N241-derived glycopeptides showed moderate diagnostic utility, particularly in early-stage HCC.

Five glycopeptides derived from two VTNC glycosylation sites (169 and 242) were identified as significantly differentially expressed across cirrhosis, early HCC, and late HCC (Figure 4). Quantitative analyses demonstrated a consistent stepwise increase in signal intensity with advancing disease stage. At glycosite 169, VTNC_169_A2G2F0S1 showed a significant elevation in mean intensity in late HCC compared with cirrhosis, while early HCC exhibited intermediate levels. A similar progressive trend was observed at glycosite 242 for VTNC_242_A2G2F0S2, where both early and late HCC groups displayed higher intensities relative to cirrhosis, with the greatest abundance detected in late HCC.

**Figure 4.**
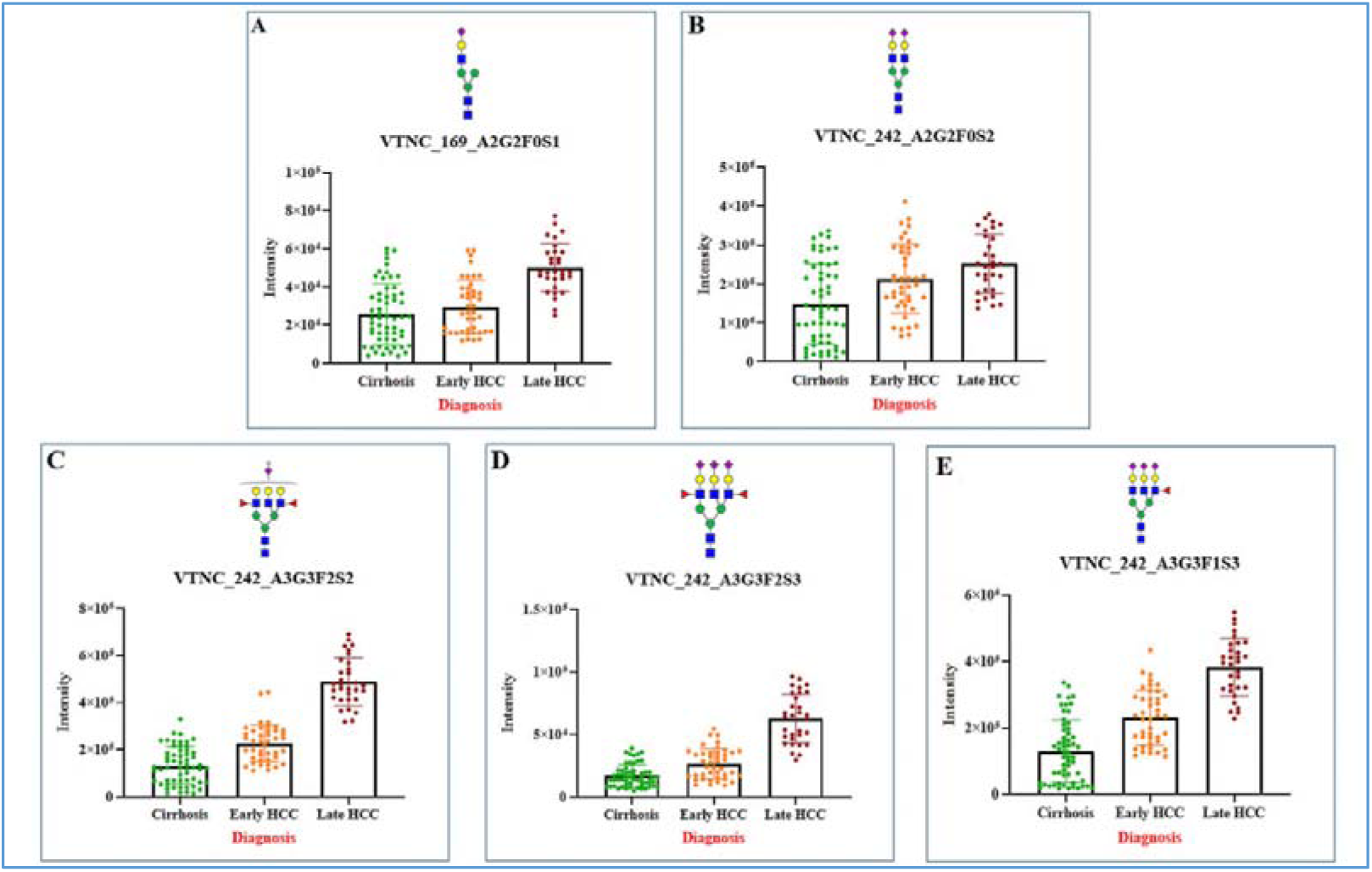
Relative abundance of VTNC glycopeptides in cirrhosis, early HCC, and late HCC. Scatter plots with overlaid bar graphs illustrate the distribution and mean intensity of individual samples for each glycopeptide across the three groups. (A) VTNC_169_A2G2F0S1; (B) VTNC_242_A2G2F0S2; (C) VTNC_242_A3G3F2S2; (D) VTNC_242_A3G3F2S3; and (E) VTNC_242_A3G3F1S3. Each dot represents an individual sample, and bars indicate the mean ± SD.

Further site-specific analysis at position 242 revealed marked increases in more complex, highly sialylated glycoforms. VTNC_242_A3G3F2S2 demonstrated a pronounced elevation in late HCC compared with both cirrhosis and early HCC. Likewise, VTNC_242_A3G3F2S3 exhibited a progressive increase across disease stages, reaching maximal levels in late HCC. The most heavily sialylated form, VTNC_242_A3G3F1S3, also showed a stepwise increase, with significant separation between cirrhosis and late HCC.

Although intra-group variability was observed across all five glycopeptides, the overall pattern consistently indicated enhanced glycopeptide intensity in HCC, particularly in late-stage disease. Notably, glycoforms characterized by increased branching and sialylation (A3G3F2S2, A3G3F2S3, and A3G3F1S3) exhibited the largest fold changes. These findings suggest that disease progression is associated not only with elevated VTNC abundance but also with increased glycan structural complexity.

ROC analysis demonstrated modest discriminatory performance for the N169 glycopeptide. The estimated AUC values and corresponding 95% CI for each individual marker are summarized in Table S3. The AUC was 0.590 (95% CI: 0.483-0.697) for all HCC with a sensitivity of 0.110 at 80% specificity, 0.624 (95% CI: 0.516-0.734) for early-stage HCC with a sensitivity of 0.190, and 0.543 (95% CI: 0.417-0.662) for late-stage HCC with no detectable sensitivity (0.000) at the same specificity threshold. Four glycopeptides were derived from glycosite N242, including one bi-antennary glycopeptide (N242_A2G2F0S2) and three tri-antennary glycopeptides (N242_A3G3F2S2, N242_A3G3F2S3, and N242_A3G3F1S3). For discrimination of all HCC cases, the corresponding AUCs were 0.643 (95% CI: 0.551-0.744), 0.632 (95% CI: 0.520-0.731), and 0.586 (95% CI: 0.485-0.685), with sensitivities of 0.219, 0.288, and 0.274 at 80% specificity, respectively. For early-stage HCC, the AUCs were 0.597 (95% CI: 0.485-0.700), 0.563 (95% CI: 0.447-0.672), and 0.534 (95% CI: 0.425-0.640), corresponding to sensitivities of 0.262, 0.119, and 0.214, respectively. Notably, improved performance was observed for late-stage HCC, with AUCs of 0.752 (95% CI: 0.652-0.850), 0.748 (95% CI: 0.645-0.845), and 0.678 (95% CI: 0.568-0.781), accompanied by sensitivities of 0.355, 0.484, and 0.323 at 80% specificity, respectively.

### 3.4. Evaluation of the Diagnostic Performance of Combinatorial Hp N-Glycopeptide Markers with AFP

To improve the discriminatory performance of AFP, we sought to identify optimal marker combinations. Considering five candidate glycopeptide markers together with age and gender, using AFP as the anchor marker, we constructed all possible 2-marker and 3-marker panels. Panels achieving the highest AUC/ROC were selected for further validation. Table S4 summarizes the performance of the best performing 2-marker panels. P-values comparing each selected panel with AFP alone were calculated using a bootstrapping approach.

All 2-marker combination models achieved AUCs greater than 0.820 for distinguishing HCC from cirrhosis, exceeding the AUC of 0.788 obtained with AFP alone. Among these, the combination of N241_A4G4F1S3 and AFP yielded the highest AUC of 0.834 (0.760,0.900). At 85% specificity, this panel achieved a sensitivity of 75.3% compared with 52.9% for AFP alone. The other three glycopeptides on site 241, N241_A2G2S1F2, N241_A3G3F1S3, and N241_A4G4F2S4, combined with AFP achieved AUCs of 0.820 (0.744,0.885), 0.816 (0.74,0.879) and 0.789 (0.717,0.867) respectively. Their sensitivity at 85% specificity were 69.9, 69.9, and 65.8%, respectively. At the site N184, the combination of the N184_A3G3F1S3 with AFP achieved an AUC of 0.826 (0.749–0.894), with a sensitivity of 75.3% at 85% specificity.

In the comparison of early-stage HCC vs cirrhosis, the AFP + N241_A4G4F1S3 panel again demonstrated the best performance, with an AUC of 0.830 (0.743,0.904), modestly higher than AFP alone (AUC 0.768; 0.672,0.860). At 85% specificity, the sensitivity of this panel was 73.8% compared with 51.4% for AFP alone.

Consistent with the findings for all-stage HCC, the combinations of AFP with the other three glycopeptides at site N241, N241_A2G2F1S2, N241_A3G3F1S3, and N241_A4G4F2S4 each achieved AUC values greater than 0.762 and demonstrated improved sensitivity at 85% specificity compared with AFP alone in distinguishing early-stage HCC from cirrhosis. At site N184, the combination of N184_A3G3F1S3 with AFP yielded an AUC of 0.872(0.791,0.938). Notably, the AFP + N184_A3G3F1S3 panel achieved the highest AUC among all evaluated glycopeptide combinations for both overall HCC and early-stage HCC detection.

### 3.5. Diagnostic Performance of Three-Marker Panel Combinations Based on Hp and VTNC

When considering three-marker panels with AFP designated as the anchor marker, optimal combinations were identified based on the highest estimated AUC values. For the comparison of all HCC vs cirrhosis, the panel comprising AFP+VTNC_169_A2G1F0S1+VTNC_242_A3G3F2S2 demonstrated the best performance, achieving an AUC of 0.859 (0.789,0.919). At 85% specificity, this model yielded a sensitivity of 76.7% and was statistically superior to AFP alone (Table S5).

In the analysis of early-stage HCCs vs cirrhosis, the combination of AFP, Age, and HP_184_A3G3F1S3 showed the highest diagnostic accuracy with an AUC of 0.904 (0.837,0.958), and a sensitivity of 76.2% at 95% specificity. An alternative panel consisting of AFP+HP_184_A3G3F1S3 and VTNC_169_A2G1F0S1 also demonstrated strong performance, with an AUC of 0.890 (0.667,0.983), and a sensitivity of 66.7% at 95% specificity. Both panels were statistically superior to AFP alone (P = 0.0226 and P=0.0393, respectively). In the late-stage analysis, the three-combination model achieved the best overall performance among all tested combinations. However, two major combinations, AFP+HP_241_A2G2F1S2+HP_241_A3G3F1S and AFP+VTNC_169_A2G2F0S1+VTNC_242_A3G3F2S2, also demonstrated strong performance, with AUCs of 0.935 (0.875–0.979) and 0.932 (0.875–0.977), respectively, and a sensitivity of 77.4% at 98% specificity.

### 3.6. SHAP-Selected Gaussian Naive Bayes Classification

A Gaussian Naive Bayes classifier was trained using seven SHAP-selected features to distinguish MASLD-associated HCC from cirrhosis. The model did not use demographic variables, including age or sex/gender. This distinction is important because several established algorithms, including GALAD, incorporate demographic variables, whereas the present classifier was restricted to molecular/glycopeptide-derived information.

In the training cohort, the confusion matrix contained 47 true negatives, 0 false positives, 2 false negatives, and 56 true positives. In the independent testing cohort, the confusion matrix contained 11 true negatives, 0 false positives, 0 false negatives, and 15 true positives. In the training cohort (n = 105), the model achieved ROC/AUC of 0.9985 and an accuracy of 98.1%. Sensitivity was 96.6%, specificity was 100.0%, precision was 100.0%, balanced accuracy was 98.3%, and F1-score was 98.2%.

In the independent testing cohort (n = 26), the model achieved ROC/AUC of 1.0000 and an accuracy of 100.0%. Sensitivity, specificity, precision, balanced accuracy, and F1-score were all 100.0% at the selected operating point. The near-perfect ROC-AUC values in both training and testing cohorts indicate excellent discrimination between MASLD-associated HCC and cirrhosis. Importantly, because the HCC cohort included both early- and late-stage cases and the model maintained very high performance, no meaningful reduction in detectability was observed for early-stage versus late-stage disease in this model. This supports the potential utility of the SHAP-selected seven-feature classifier for early detection, although stage-specific validation should be reported if separate early- and late-stage confusion matrices become available.

The choice to maintain a held-out test cohort rather than relying only on cross-validation is important in this context. Although cross-validation is often expected in small biomarker studies, the strong apparent separation of the SHAP-selected features allowed independent testing without undermining model training. This design provides a clearer demonstration that the classifier was not simply optimized on the same samples used for evaluation.

### 3.7. Comparison with AFP, Logistic-Regression Panels, and Related Glycopeptide-Based Methods

To contextualize the machine-learning classifier, its performance was compared with AFP alone, the best logistic-regression panels from the present study, and related glycopeptide-based HCC biomarker approaches reported in prior studies. Because the Naive Bayes results were obtained from a patient-level train/test split whereas several comparator values were estimated from ROC analyses or logistic-regression panels in the full cohort, the comparison should be interpreted as exploratory rather than as a formal head-to-head validation.

The Gaussian Naive Bayes classifier substantially exceeded the AUC values observed for AFP alone and for the best logistic-regression panels in this manuscript. The test-set AUC of 1.0000 and accuracy of 100.0% suggest that the seven SHAP-selected molecular features captured a highly discriminative glycopeptide signature for MASLD-associated HCC. The exclusion of demographic variables is also a practical advantage. Demographic variables can improve apparent classification in retrospective datasets but may introduce limitations if the validation cohort differs in age, sex distribution, ethnicity, or recruitment setting. A biomarker model that does not require demographic covariates may therefore be easier to evaluate prospectively and less dependent on cohort matching.

Because the model maintained near-perfect discrimination despite inclusion of both early- and late-stage HCC cases, the classifier did not show evidence of poorer performance for early-stage disease. This is an important distinction from several individual glycopeptides and AFP-based models, where diagnostic performance often differs between early- and late-stage HCC. Despite the strong performance, these results should be interpreted with appropriate caution because the testing cohort contained 26 patients. External validation in an independent MASLD-HCC cohort and stage-stratified testing will be required to confirm generalizability and to determine whether the model remains equally accurate for early-stage disease in broader clinical settings.

Relative to these published approaches, the SHAP-selected Gaussian Naive Bayes classifier in the present manuscript showed higher apparent discrimination while avoiding demographic inputs. Because both early- and late-stage HCC cases were included and the ROC-AUC remained near-perfect, the model did not show evidence of reduced early-stage detection performance in this dataset. This comparison should nevertheless be described as exploratory because external validation has not yet been performed.

## 4. CONCLUSIONS

In this study we employed NanoLC-PRM-MS/MS to analyze glycopeptides directly from clinical serum samples for the identification of biomarkers for early detection of HCC. Unlike previous approaches, this methodology eliminates the need for isolation of the target marker proteins, thereby streamlining the workflow and enhancing its suitability for clinical application.

We focused on the protein markers Hp and VTNC by examining their glycopeptide profiles in a cohort of MASLD patient samples collected from hospital in Holhot, China. Glycopeptides from these proteins were enriched and analyzed by Mass Spectrometry using the selectivity of PRM-MS. Through this approach We identified several marker combinations that, when used in conjunction with AFP, substantially improved the AUC for HCC detection across both early-stage and all-stage samples

Using a three-marker panel with AFP as the anchor, the optimal combination for distinguishing all HCC vs cirrhosis was AFP+VTNC_169_A2G1F0S1+VTNC_242_A3G3F2S2, achieving an estimated AUC of 0.859 (0.789,0.919), with 76.7% sensitivity at 90% specificity. For early-stage HCC vs cirrhosis, the combination of AFP+HP_184_A3G3F1S3+VTNC_169_A2G1F0S1, resulted in an AUC of 0.837 (0.761,0.896), with 60.3% sensitivity at 95% specificity. These panels represent a marked improvement over AFP alone, which yielded an AUC of 0.935 and a sensitivity of 77.4% at 100% specificity. Overall, these findings demonstrate that targeted glycopeptide analysis via NanoLC-PRM-MS/MS can enhance the early detection of HCC and provide a clinically practical approach for biomarker-driven diagnostics.

The addition of a seven-feature SHAP-selected Gaussian Naive Bayes classifier further strengthened diagnostic performance, achieving ROC-AUC values of 0.9985 in training and 1.0000 in independent testing, with corresponding accuracies of 98.1% and 100.0%. Importantly, this model did not use demographic variables, which may reduce cohort-specific bias and simplify prospective validation. These results suggest that the selected molecular/glycopeptide signature can detect MASLD-associated HCC with high accuracy without an apparent loss of performance for early-stage disease, although external validation remains necessary.

## DATA AVAILABILITY

The MS raw data generated using the Orbitrap NanoLC-HCD-PRM-MS/MS workflow are available from the corresponding author upon reasonable request.

## SUPPORTING INFORMATION

The following supporting information is available free of charge at ACS website http://pubs.acs.org

**Table-S1:** Clinical Information of 131 Samples and Intensities of Twenty Target Glycoforms (xlsx).

**Table-S2:** List of Precursors Used for PRM Window Setting.

**Table-S3:** Diagnostic Performance (AUC) of Individual Markers at Different Disease Stages.

**Table-S4:** Diagnostic Performance of Combinatorial Hp N-Glycopeptide Markers with AFP at Sites N241and N184 for all HCC, Early HCC, and Late HCC.

**Table-S5:** Diagnostic Performance of Combinatorial Three-Marker Panel Based on Hp and VTNC.

## Supporting information

Supplemental table -1 Clinical Information

## ACKNOWLEDGMENTS

We gratefully acknowledge partial support by the National Cancer Institute for this work through grant R01-CA160254-11 (D.M.L.). Additionally, D.M.L. acknowledges support from the Maud T. Lane Professorship.

## AUTHOR CONTRIBUTIONS

**Y.L., C.V., and D.M.L.** conceptualized the study and led the study design and data analysis**. J.D.** and **S.Y.** performed the statistical analyses. **N.Y.L.** conducted the machine learning analysis. **C.V.** and **Y.L.** drafted and revised the manuscript. All authors reviewed and approved the final manuscript.

