## Supplementary Information for "Predicting Early MASLD-HCC from Serum N-Glycomics: A SHAP-Interpreted Gaussian Naive Bayes Model Built on nLC-HCD-PRM-MS/MS Profiling"

† Equal first authorship

*Corresponding Author

David M. Lubman

Department of Surgery, University of Michigan Medical Center

Ann Arbor, Michigan 48109, United States

orcid.org/0000-0001-7731-0232

**Table-S1:** Clinical Information of 131 Samples and Intensities of Twenty Target Glycoforms (xlsx)

**Table-S2: List of Precursors Used for PRM Window Setting**

| ***Protein name\|ID\|Abbr_Glycosite*** | ***Glycopeptides*** | ***m/z*** | ***charge*** | ***t start (min)*** | ***t stop (min)*** |
|---|---|---|---|---|---|
| *Vitronectin \| P04004 \| VTNC_242* | A4G4F1S3 | 1258.92 | 5 | 60 | 65.0 |
| *NISDGFDGIPDNVDAALALPAHSYSGR* | A4G4F0S4 | 1258.72 | 5 | 60 | 65.0 |
| | A3G3F2S3 | 1482.12 | 4 | 48.3 | 53.3 |
| | A3G3F1S3 | 1445.62 | 4 | 48.5 | 53.5 |
| | A3G3F4S1 | 1127.88 | 5 | 48.7 | 53.7 |
| | A3G3F2S2 | 1127.67 | 5 | 48.6 | 53.6 |
| | A3G3F3S1 | 1373.08 | 4 | 43.1 | 48.1 |
| | A2G2F1S2 | 1281.54 | 4 | 43.3 | 48.3 |
| | A2G2F2S1 | 1245.28 | 4 | 43. | 48.0 |
| | A2G2F0S2 | 1244.52 | 4 | 43. | 48.0 |
| *Vitronectin \| P04004 \| VTNC_169* | A2G2F0S2 | 1039.42 | 3 | 39.5 | 44.5 |
| *NGSLFAFR* | A2G2F0S1 | 943.06 | 3 | 39.5 | 44.5 |
| *Vitronectin \| P04004 \| VTNC_86* | A2G2F0S2 | 1529.32 | 3 | 26.6 | 31.6 |
| *NNATVHEQVGGPSLTSDLQAQSK* | A3G3F2S2 | 1311.55 | 4 | 29.7 | 34.7 |
| | A2G2F1S2 | 1183.76 | 4 | 26.5 | 31.5 |
| *Haptoglobin \| P00738 \| Hp_184* | A2G2F1S2 | 1022.93 | 4 | 20.4 | 25.4 |
| *MVSHHJLTTGATLINEQWLLTTAK* | A2G2F1S2 | 1026.93 | 4 | 20.3 | 25.3 |
| | A2G2F1S2 | 1363.57 | 3 | 20.4 | 25.4 |
| | A2G2F1S2 | 1368.90 | 3 | 20.3 | 25.3 |
| | A3G3F1S2 | 1114.21 | 4 | 20.7 | 25.7 |
| | A3G3F1S2 | 1485.28 | 3 | 20.7 | 25.7 |
| | A3G3F1S3 | 1186.98 | 4 | 23.9 | 28.9 |
| | A3G3F1S3 | 1190.98 | 4 | 23.8 | 28.8 |
| | A3G3F1S3 | 1582.31 | 3 | 23.9 | 28.9 |
| | A3G3F1S3 | 1587.64 | 3 | 23.8 | 28.8 |
| | A3G3F2S2 | 1150.73 | 4 | 25.8 | 30.8 |
| | A3G3F2S2 | 1154.73 | 4 | 25.8 | 30.8 |
| | A3G3F2S3 | 1223.49 | 4 | 26.5 | 31.5 |
| | A3G3F2S3 | 1227.49 | 4 | 26.5 | 31.5 |
| | A3G3F2S3 | 1630.99 | 3 | 26.5 | 31.5 |
| | A3G3F2S3 | 1634.99 | 3 | 26.5 | 31.5 |
| *Haptoglobin \| P00738 \| Hp_241* | A2G2F1S2 | 1037.21 | 4 | 31.7 | 36.7 |
| *VVLHPnYSQVDIGLIK* | A2G2F1S2 | 1382.61 | 3 | 31.7 | 36.7 |
| | A3G3F1S1 | 1407.30 | 3 | 26.7 | 31.7 |
| | A3G3F1S2 | 1504.32 | 3 | 31.6 | 36.6 |
| | A3G3F1S3 | 1201.27 | 4 | 35.9 | 40.9 |
| | A3G3F1S3 | 1601.36 | 3 | 35.9 | 40.9 |
| | A3G3F2S2 | 1165.01 | 4 | 35.4 | 41.4 |
| | A3G3FS3 | 1164.76 | 4 | 35.4 | 41.4 |
| | A3G3F2S3 | 1237.78 | 4 | 35.1 | 41.9 |
| | A3G3F2S3 | 1650.05 | 3 | 35.1 | 41.9 |
| | A4G4F1S3 | 1292.55 | 4 | 36.5 | 41.5 |
| | A4G4F1S3 | 1723.07 | 3 | 36.5 | 41.5 |
| | A4G4F1S4 | 1365.33 | 4 | 36.9 | 41.9 |
| | A4G4F1S4 | 1820.11 | 3 | 36.9 | 41.9 |
| | A4G4F2S4 | 1401.84 | 4 | 39.9 | 49.9 |
| | A4G4F2S4 | 1868.79 | 3 | 39.9 | 49.9 |

**Table – S3: Diagnostic Performance (AUC) of Individual Markers at Different Disease Stages**

| *Markers* | *Diagnosis* | *AUC (CI:95%)* | *Sens / Spec 80 %* | *P-value* |
|---|---|---|---|---|
| *AFP* | All_HCC | 0.788 (0.711,0.861) | 0.679 | <0.0001 |
| *Age* | All_HCC | 0.690 (0.601,0.777) | 0.458 | 0.0002 |
| *HP_241_ A3G3F1S3* | All_HCC | 0.672 (0.582,0.756) | 0.411 | 0.0007 |
| *HP_241_ A4G4F1S3* | All_HCC | 0.662 (0.565,0.754) | 0.397 | 0.0015 |
| *HP_241_ A2G2F1S2* | All_HCC | 0.650 (0.551,0.746) | 0.301 | 0.0032 |
| *VTNC_242_ A3G3F2S2* | All_HCC | 0.643 (0.551,0.744) | 0.219 | 0.0049 |
| *HP_184_ A3G3F1S3* | All_HCC | 0.637 (0.528,0.731) | 0.493 | 0.0075 |
| *VTNC_N242_ A3G3F1S3* | All_HCC | 0.632 (0.527,0.731) | 0.288 | 0.0099 |
| *VTNC_242_ A2G2F0S1* | All_HCC | 0.603 (0.495,0.702) | 0.247 | 0.0431 |
| *Gender* | All_HCC | 0.596 (0.516,0.686) | 0.318 | 0.0277 |
| *VTNC_169_ A2G2F0S1* | All_HCC | 0.590 (0.483,0.697) | 0.110 | 0.0787 |
| *VTNC_242_ A3G3F2S3* | All_HCC | 0.586 (0.485,0.685) | 0.274 | 0.0930 |
| *VTNC_242_ A2G2F0S2* | All_HCC | 0.554 (0.453,0.660) | 0.027 | 0.2939 |
| *HP_241_ A4G4F2S4* | All_HCC | 0.552 (0.444,0.656) | 0.260 | 0.3047 |
| *AFP* | Early_HCC | 0.768 (0.672,0.860) | 0.667 | <0.0001 |
| *HP_184_A3G3F1S3* | Early_HCC | 0.766 (0.665,0.863) | 0.690 | <0.0001 |
| *Age* | Early_HCC | 0.724 (0.620,0.820) | 0.510 | 0.0001 |
| *HP_241_A4G4F1S3* | Early_HCC | 0.715 (0.609,0.814) | 0.548 | 0.0003 |
| *HP_241_ A3G3F1S3* | Early_HCC | 0.624 (0.514,0.725) | 0.357 | 0.0352 |
| *VTNC_169_ A2G2F0S1* | Early_HCC | 0.624 (0.516,0.734) | 0.190 | 0.0346 |
| *HP_241_ A4G4F2S4* | Early_HCC | 0.620 (0.509,0.723) | 0.333 | 0.0411 |
| *Gender* | Early_HCC | 0.610 (0.516,0.701) | 0.334 | 0.0289 |
| *VTNC_242_ A3G3F1S3* | Early_HCC | 0.597 (0.485,0.700) | 0.262 | 0.0986 |
| *VTNC_242_ A3G3F2S2* | Early_HCC | 0.563 (0.447,0.672) | 0.119 | 0.2837 |
| *HP_241_ A2G2F1S2* | Early_HCC | 0.562 (0.455,0.668) | 0.167 | 0.2900 |
| *VTNC_242_ A3G3F2S3* | Early_HCC | 0.534 (0.425,0.640) | 0.214 | 0.5645 |
| *VTNC_242_ A2G2F0S2* | Early_HCC | 0.513 (0.406,0.628) | 0.048 | 0.8313 |
| *VTNC_242_ A2G2F0S1* | Early_HCC | 0.462 (0.348,0.586) | 0.000 | 0.5183 |
| *AFP* | Late_HCC | 0.815 (0.717,0.906) | 0.697 | <0.0001 |
| *HP_241_ A2G2F1S2* | Late_HCC | 0.769 (0.653,0.865) | 0.484 | <0.0001 |
| *VTNC_242_A3G3F2S2* | Late_HCC | 0.752 (0.652,0.850) | 0.355 | 0.0001 |
| *VTNC_242_ A3G3F2S3* | Late_HCC | 0.748 (0.645,0.845) | 0.484 | 0.0001 |
| *HP_241_ A3G3F1S3* | Late_HCC | 0.737 (0.625,0.837) | 0.484 | 0.0002 |
| *VTNC_242_ A2G2F0S1* | Late_HCC | 0.691 (0.581,0.790) | 0.355 | 0.0031 |
| *VTNC_N242_ A3G3F1S3* | Late_HCC | 0.678 (0.568,0.781) | 0.323 | 0.0059 |
| *Age* | Late_HCC | 0.644 (0.519,0.759) | 0.387 | 0.0261 |
| *VTNC_242_A2G2F0S2* | Late_HCC | 0.609 (0.488,0.728) | 0.000 | 0.0923 |
| *HP_241_ A4G4F1S3* | Late_HCC | 0.590 (0.472,0.708) | 0.194 | 0.1643 |
| *Gender* | Late_HCC | 0.578 (0.472,0.677) | 0.295 | 0.1520 |
| *VTNC_169_ A2G2F0S1* | Late_HCC | 0.543 (0.417,0.662) | 0.000 | 0.5100 |
| *HP_184_ A3G3F1S3* | Late_HCC | 0.539 (0.413,0.668) | 0.323 | 0.5495 |
| *HP_241_ A4G4F2S4* | Late_HCC | 0.539 (0.412,0.662) | 0.258 | 0.5438 |

ROC analysis was performed to evaluate the ability of each individual marker to discriminate HCC cases from controls in the overall cohort (All_HCC) and by stage (Early_HCC and Late_HCC). The ROC/AUC, with corresponding 95% CI is reported for each marker. Sensitivity values are presented at a fixed specificity of 80% (Sens / Spec 80%). Statistical significance of each AUC compared with the null hypothesis (AUC = 0.5) was assessed using two-sided tests, and the corresponding p-values are shown. Markers include the conventional clinical variable AFP, demographic factors (age and gender), and site-specific glycopeptide features derived from HP and VTNC. Higher AUC values indicate better diagnostic discrimination.

**Table S4. Diagnostic Performance of Combinatorial Hp N-Glycopeptide Markers with AFP at Sites N241and N184 for all HCC, Early HCC, and Late HCC.**

| *Marker HP_241* | *All HCC vs Cirrhosis* | | | | *Early-Stage vs Cirrhosis* | | | | *Late*-Stage *vs Cirrhosis* | | | |
|---|---|---|---|---|---|---|---|---|---|---|---|---|
| | *AUC* | *95% CI* | *P-Val.* | *Sens/Spec* | *AUC* | *95% CI* | *P-Val.* | *Sens/Spec* | *AUC* | *95% CI* | *P-Val.* | *Sens/Spec* |
| *AFP+ A4G4F1S3* | 0.834 | (0.760-0.900) | 0.3644 | 0.753/0.793 | 0.830 | (0.743-0.904) | 0.3402 | 0.738/0.793 | 0.844 | (0.753-0.926) | 0.6834 | 0.774/0.828 |
| *AFP+ A2G2F1S2* | 0.820 | (0.744-0.885) | 0.5493 | 0.699/0.828 | 0.785 | (0.694-0.874) | 0.8001 | 0.69/ 0.81 | 0.877 | (0.791-0.944) | 0.3537 | 0.613/1 |
| *AFP+ A3G3F1S3* | 0.816 | (0.74-0.879) | 0.5943 | 0.699/0.828 | 0.787 | (0.69-0.878) | 0.7759 | 0.643/0.845 | 0.864 | (0.771,0.941) | 0.4713 | 0.645/ 0.948 |
| *AFP+A4G4F2S4* | 0.798 | (0.717-0.867) | 0.8564 | 0.658/0.862 | 0.802 | (0.702-0.886) | 0.6094 | 0.738/0.81 | 0.816 | (0.716-0.909) | 0.9939 | 0.548/0.983 |
| *Marker HP_184* | *All HCC vs Cirrhosis* | | | | *Early-Stage vs Cirrhosis* | | | | *Late-Stage vs Cirrhosis* | | | |
| | *AUC* | *95% CI* | *P-Val.* | *Sens/Spec* | *AUC* | *95% CI* | *P-Val.* | *Sens/Spec* | *AUC* | *95% CI* | *P-Val.* | *Sens/Spec* |
| *AFP+ A3G3F1S3* | 0.826 | (0.749,0.894) | 0.4759 | 0.753/0.828 | 0.872 | (0.791,0.938) | 0.0927 | 0.738/ 0.862 | 0.843 | (0.75-0.93) | 0.6902 | 0.548/1 |

**Table S5. Diagnostic Performance of Combinatorial Three-Marker Panel Based on Hp and VTNC**

| *Marker panel* | *All HCC vs Cirrhosis* | | | | *Early-Stage vs Cirrhosis* | | | | *Late*-Stage *vs Cirrhosis* | | | |
|---|---|---|---|---|---|---|---|---|---|---|---|---|
| | *AUC* | *95% CI* | *P-Val.* | *Sens/ Spec* | *AUC* | *95% CI* | *P-Val.* | *Sens/ Spec* | *AUC* | *95% CI* | *P-Val.* | *Sens/ Spec* |
| *AFP+VTNC_169_ A2G2F0S1+VTNC_242_A3G3F2S 2* | 0.859 | (0.789,0.919) | 0.1557 | 0.767/ 0.897 | 0.818 | (0.727-0.902) | 0.4496 | 0.69/0. 897 | 0.932 | (0.875-0.977) | 0.0477 | 0.839/ 0.914 |
| *AFP+Age+HP_184_ A3G3F1S3* | 0.847 | (0.772-0.907) | 0.2535 | 0.685/ 0.931 | 0.904 | (0.837,0.958) | 0.0226 | 0.762/ 0.914 | 0.854 | (0.765-0.928) | 0.5639 | 0.71/ 0.845 |
| *AFP+HP_184_ A3G3F1S3+VTNC_169_ A2G2F0S1* | 0.837 | (0.761-0.896) | 0.3468 | 0.603/ 0.931 | 0.89 | (0.667/0.983) | 0.0392 | 0.667/ 0.983 | 0.848 | (0.757-0.926) | 0.6412 | 0.548/ 1 |
| *AFP+HP_241_A2G2F1S2+HP_2 41_A3G3F1S3* | 0.856 | (0.791-0.916) | 0.182 | 0.712/ 0.897 | 0.806 | (0.709-0.892) | 0.5743 | 0.738/ 0.862 | 0.935 | (0.875,0.979) | 0.0401 | 0.774/ 0.983 |
| *AFP+VTNC_169_A2G2F0S1+VT NC_242_A3G3F2S2* | | | | | | | | | 0.932 | (0.875,0.977) | 0.0477 | |
